# Autoantibodies against IL-1-receptor-antagonist in multisystem inflammatory syndrome in children

**DOI:** 10.1101/2021.09.08.21263027

**Authors:** Jochen Pfeifer, Bernhard Thurner, Christoph Kessel, Natalie Fadle, Evi Regitz, Marie-Christin Hoffmann, Igor Kos, Klaus-Dieter Preuss, Yvan Fischer, Klaus Roemer, Stefan Lohse, Kristina Heyne, Marie-Claire Detemple, Michael Fedlmeier, Hendrik Juenger, Harald Sauer, Sascha Meyer, Tilman Rohrer, Helmut Wittkowski, Katja Masjosthusmann, Sören L. Becker, Sigrun Smola, Moritz Bewarder, Michael Boehm, Jordi Anton, Rosa Maria Pino-Ramirez, Hashim Abdul-Khaliq, Dirk Foell, Lorenz Thurner

## Abstract

Multisystem inflammatory syndrome in children (MIS-C or PIMS) is a rare but serious complication after an infection with SARS-CoV-2. A possible involvement of pathogenetically relevant autoantibodies has been discussed. Recently neutralizing autoantibodies against anti-inflammatory receptor antagonists progranulin (PGRN) and IL-1-receptor antagonist (IL-1-Ra) were discovered in adult patients with critical COVID-19.

Plasma of an index case with severe PIMS/MIS-C was analyzed for autoantibodies against IL-1-Ra and PGRN. The study was extended by a case series of 12 additional patients. In addition to ELISA for of antibodies, IL-1-Ra plasma levels were determined and IL-1-Ra was analyzed by Western-blot and isoelectric focusing. Functional activity of the autoantibodies was examined in vitro with IL-1ß reporter assays.

Antibodies against IL-1-Ra could be detected in 10 of 13 (76.9%) patients with PIMS/MIS-C, but not in controls. In contrast to critical COVID-19 in adults, no IL-1-Ra antibodies of the IgM class were detected in PIMS/MIS-C. IL-1-Ra-antibodies exclusively belonged to IgG1. No antibodies directed against PGRN were detected. Western blots and ELISA showed a concomitant reduction of free IL-1-Ra plasma levels in the presence of IL-1-Ra-antibodies. The antibodies inhibited IL-1-Ra function in IL-1ß reporter cell assays. Notably, an additional, hyperphosphorylated, transiently occurring atypical isoform of IL-1-Ra was observed in all IL-1-Ra autoantibody-positive patients.

To conclude, IL-1-Ra autoantibodies were observed in high frequency in children with PIMS/MIS-C. They may represent a diagnostic and pathophysiologically relevant marker for PIMS/MIS-C. Their generation is likely to be triggered by an atypical, hyperphosphorylated isoform of IL-1-Ra.

## Introduction

SARS-CoV-2 infection typically takes a mild or asymptomatic course in children. Pediatric inflammatory multisystem syndrome (PIMS) or multisystem inflammatory syndrome in children (MIS-C) is a rare but serious complication that usually occurs after an interval of 2 to 6 weeks after infection with SARS-CoV-2 (*1*)(*2*)(*3*). All affected children have persistent fever, other clinical features may vary. Acute abdominal pain, diarrhea or vomiting, muscle pain, headache and fatigue are frequently. Bilateral conjunctival injection (hyperemia) and exanthema, swollen hands and feet and a “strawberry tongue” resembling Kawasaki’s disease have been described in a high percentage of patients with PIMS/MIS-C. Affection of the cardiovascular system with myocardial dysfunction, arterial hypotension or systemic shock all have been regularly observed. Myocarditis, myocarditis, pericarditis and involvement of the valves and coronary arteries (dilatation or aneursyma) may develop. Laboratory studies regularly show intensive inflammation with elevated serum levels of C-reactive protein, procalcitonin, ferritin, cardiac involvement with elevated troponin and N-terminal pro-B-type natriuretic peptide, as well as hyponatremia, markers of coagulopathy (elevated D-dimers, prolonged prothrombin time and partial thromboplastin time) and hematologic abnormalities (anemia, lymphocytopenia and thrombocytopenia or thrombocytosis). Most affected children will need intensive care due to multiorgan failure and shock (*4*)(*5*)(*6*)(*7*)(*8*)(*9*).

Little is known about the pathogenesis of PIMS/MIS-C. It has been suggested that the formation of pathogenetically relevant autoantibodies contributes to a hyperinflammatory state (*10*). This is based on the observations that i) there is a interval between the infection with SARS-CoV-2 and the manifestation of the disease in the majority of cases and ii) the patients typically respond to intravenous immunoglobulins (IVIGs) and glucocorticoids. Besides stabilization with fluids, vasoactive or inotropic drugs, oxygen, acetylsalicylic acid in cases with cardiac involvement and thrombocytosis, anticoagulation in severe impairment of the left ventricle, the administration of IVIGs and steroids are presently standard-of-care. The American college of Rheumatology recommends high dose anakinra for IVIG-refractory PIMS/MIS-C, a recombinant modified human IL-1-Ra (*11*)(*12*)(*13*).

Recently, in adult patients with severe and critically ill with COVID-19, we discovered neutralizing autoantibodies against progranulin (PGRN), an anti-inflammatory ligand of the pro-inflammatory receptors TNFR1/TNFR2 and DR3, and a direct receptor antagonist of TNF-α as well as TL1a. In these patients we also discovered autoantibodies against interleukin-1 receptor antagonist (IL-1-Ra), the ligand of the pro-inflammatory IL-1 receptor and antagonist of IL-1α and Il-1β.

In addition, we found evidence that the autoantibodies probably have been evoked by a preceding and transient hyperphosphorylation of the antigens. The antibodies belonged to a broad range of Ig classes including IgM and several IgG subclasses, indicating recent immune responses (*14*). PGRN-antibodies (Abs) and the immunogenic pSer81 PGRN isoform have been initially described in systemic primary vasculitides (*15*)(*16*). As critical COVID-19 can partially resemble hyperinflammatory syndromes and has similarities to hemophagocytic lymphohistiocytosis (HLH), autoinflammatory syndromes (*17*), Kawasaki disease (*9*), and vasculitides (*18*) we performed a targeted screen for these autoantibodies and posttranslationally altered isoforms of the corresponding autoantigens, in a young girl affected with PIMS/MIS-C.

## Index Case

A formerly healthy girl, with preexisting mild allergic rhinitis due to pollen allergy, in the age group between 6 and 10 years was admitted with a history of persisting high fever for 5 days, abdominal pain, vomiting, diarrhea, slight bilateral (non-purulent) conjunctivitis and exanthema. The general condition was poor. Six weeks before admission she had a PCR-confirmed mild infection with Sars-CoV-2 detected due to another positive case in the family. On admission, echocardiography showed a reduced left ventricular shortening fraction (25%), mitral regurgitation, and progressive dilatation of the right coronary artery occurred (maximal ostial diameter 6 mm, Z-score +8,8), interestingly with a congenitally single coronary ostium. Bilateral pleural effusions and ascites were present, as well as tachycardia, tachypnoea and arterial hypotension. Laboratory results on admission showed elevated C-reactive protein of 177 mg/l, elevated NT-pro-BNP of 2,069 pg/ml (in the course max. 52,000 pg/ml), hyponatremia of 129 mmol/l and hypokalemia of 3.0 mml/l. Antibodies against Sars-CoV-2 were present.

The diagnosis of PIMS/MIS-C was suspected. Differential diagnoses were Kawasaki disease, macrophage activation syndrome, bacterial sepsis, toxic shock syndrome and other inflammatory or infectious diseases. Treatment with volume resuscitation, high-dose acetylsalicylic acid (30 mg/kg/d), IVIG (2 g/kg) and antibiotic treatment with piperacillin / tazobactam and metronidazole was initiated. Due to development of shock symptoms she was transferred to pediatric ICU requiring inotropic support (epinephrine, milrinone, levosimendan). The therapy included non-invasive ventilation (max. FiO2 0.5), bilateral pleural and ascites drainages, fluid and electrolyte management, buffering, erythrocyte transfusion, substitution of antithrombin-III, albumin and fresh-frozen-plasma. After exclusion of a bacterial or viral infection, pulse treatment with intravenous methylprednisolone 30 mg/kg/d for 5 days, followed by prednisolone 2 mg/kg/d (with gradual tapering) was commenced on the second day. A crucial improvement was seen only after the onset of glucocorticoid treatment. 72 h after defervescence the dose of acetylic salicylic acid was reduced to 100 mg per day. The patient underwent a therapy with enalaprile, metoprolole and decongestion with diuretics. On day 16 the girl could be discharged. In follow-up, echocardiographic findings and blood parameters were normalized (restitutio ad integrum).

**Table 1:**
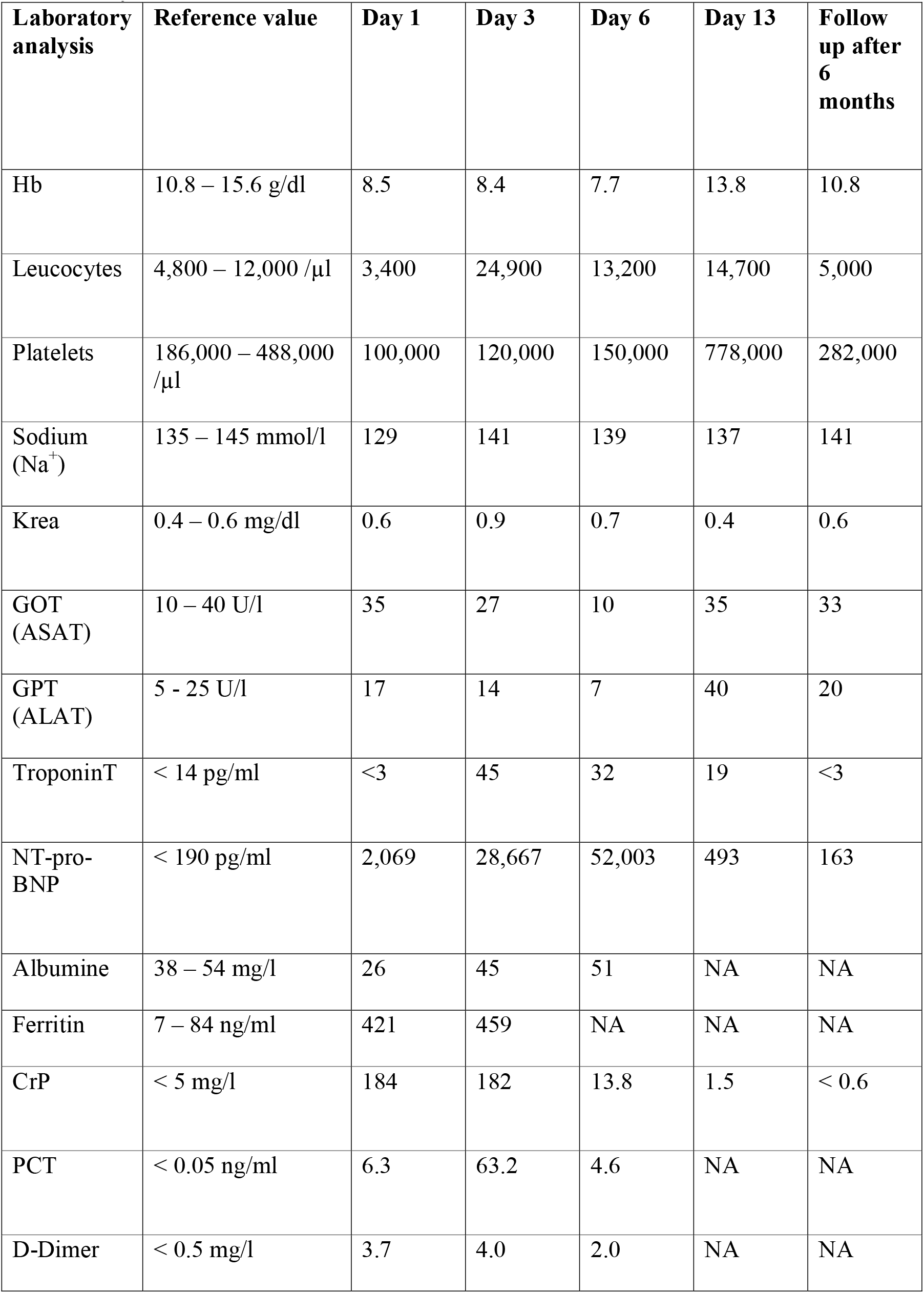
Laboratory Characteristics of the Index Patient.

## RESULTS

Plasma samples of the index case and of a case series of further 12 children with PIMS/MIS-C were analyzed. All patients survived PIMS/MIS-C.

**Table 2:**
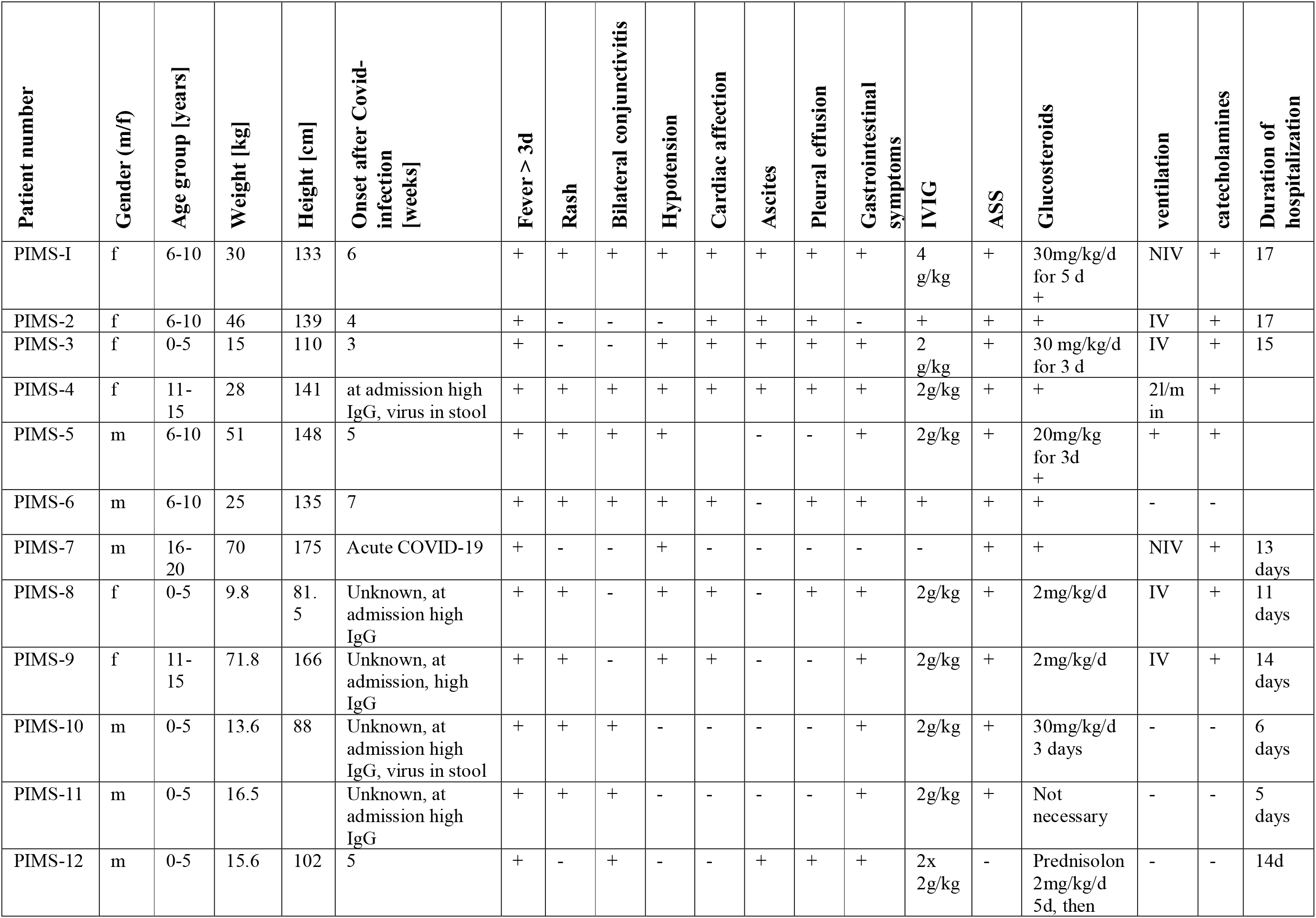
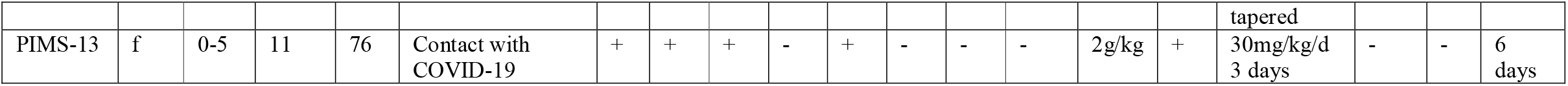
Patient characteristics.

### ELISA for IL-1-Ra-Antibodies

In the index case IL-1-Ra-Abs, but no PGRN-Abs were detected. The titers of IL-1-Ra-Abs at presentation were 1:800 (Fig. 1). IL-1-Ra-Abs belonged to IgG subclass 1 (Fig. 1), no IL-1-Ra-Abs of IgM class were observed. Three months later the titer had fallen off to 1:400 and no IL-1-Ra-Abs could be detected seven month later.

**Figure 1:**
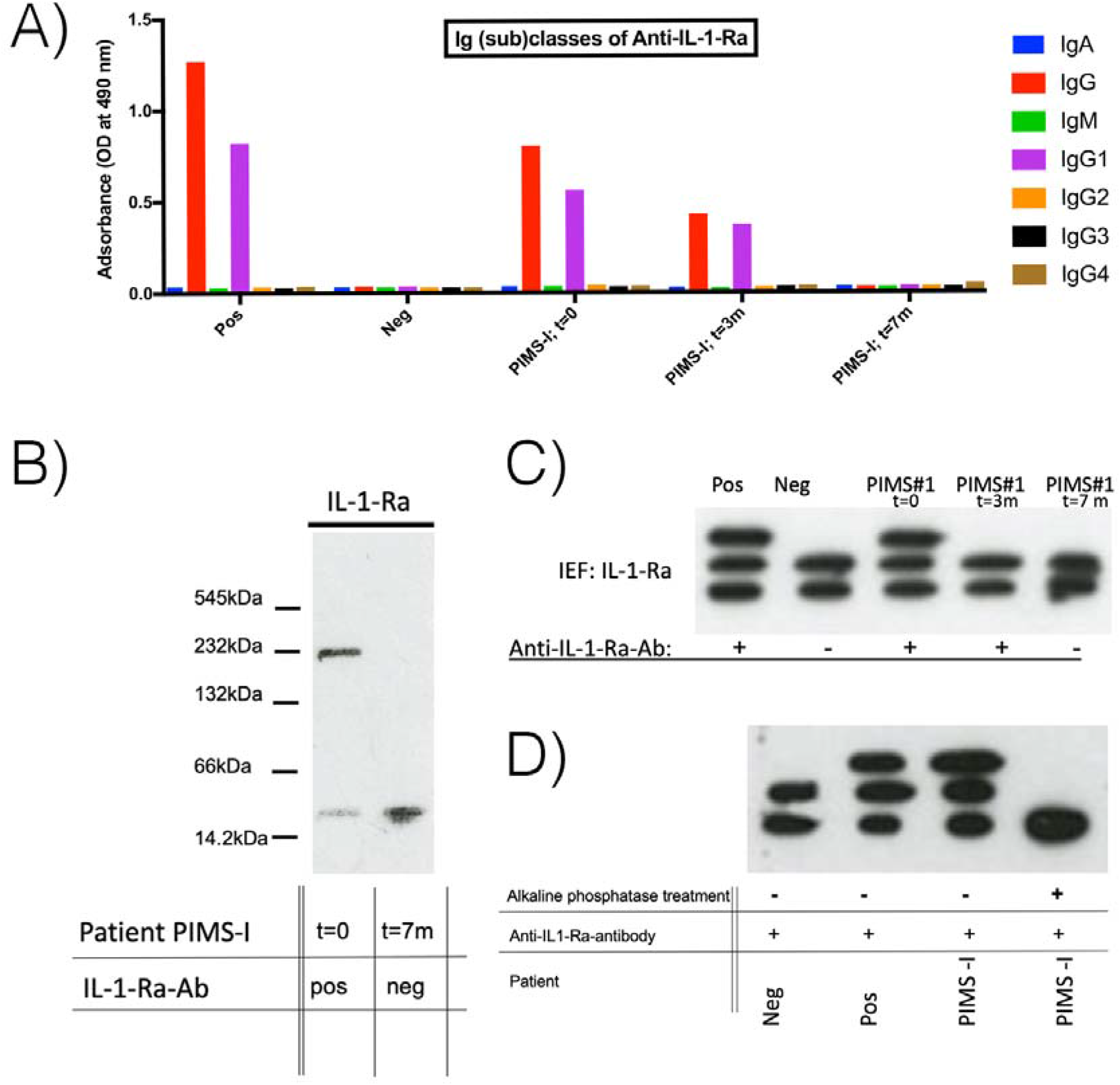
A) ELISA: Anti-IL-1-Ra-Abs and the Ig classes at the acute inflammatory phase, 3 and 7 months later of the index case. B) Native Western blot of IL-1-Ra from plasma of the index case showed a strongly reduced band of free Il-1-Ra, but an additional band of IgG-bound IL-1-Ra during the acute inflammatory phase compared to a sample 7 month later, when the patient was seronegative for IL-1-Ra-Abs. C) Isoelectric focusing (IEF) of IL-1-Ra from samples at acute inflammatory phase of index case and in the follow-up. IEF showed an additional, more negatively charged third band of IL-1-Ra at the point of time at presentation. In the further course of time this hyperphosphorylation of IL-1-Ra was not detectable 3 or 7 months after the inflammatory multisystem syndrome. D) Alkaline phosphatase treatment of IL-1-Ra of the index case. Pretreatment with alkaline phosphatase before IEF led to the disappearance of both the normally occurring second and the atypical additional third isoform IL-1-Ra isoform, confirming a hyperphosphorylation.

In the series of further 12 patients with PIMS/MIS-C, in 9 patients IL-1-Ra-Abs were detected with titers ranging between 1:200 to 1:800. All IL-1-Ra-Abs belonged exclusively to IgG1 in all patients. Autoantibodies against progranulin were not detectable in any of these patients with PIMS/MIS-C. In contrast, IL-1-Ra-Abs were not detectable in 6 patients with Kawasaki’s disease. Moreover, IL-1-Ra-Abs were not found in plasma or serum samples of 33 children of the control group.

### Atypical isoform of IL-1-Ra

IEF performed on total protein from plasma of the index case at the time of presentation, revealed the presence of an additional, more negatively charged third band of IL-1-Ra. Pretreatment with alkaline phosphatase before IEF led to the disappearance of both the normally occurring second and the atypical additional third isoform IL-1-Ra isoform, indicating a hyperphosphorylation (Fig. 1). In the further course, the hyperphosphorylated third signal of IL-1-Ra was no longer detectable at 3 or 7 months after the onset of the inflammatory multisystem syndrome in plasma of the index-patient.

The hyperphosphorylated IL-1-Ra isoform was observed in all 10 patients with IL-1-Ra-Abs but not in any of the controls (Fig. 2).

**Figure 2:**
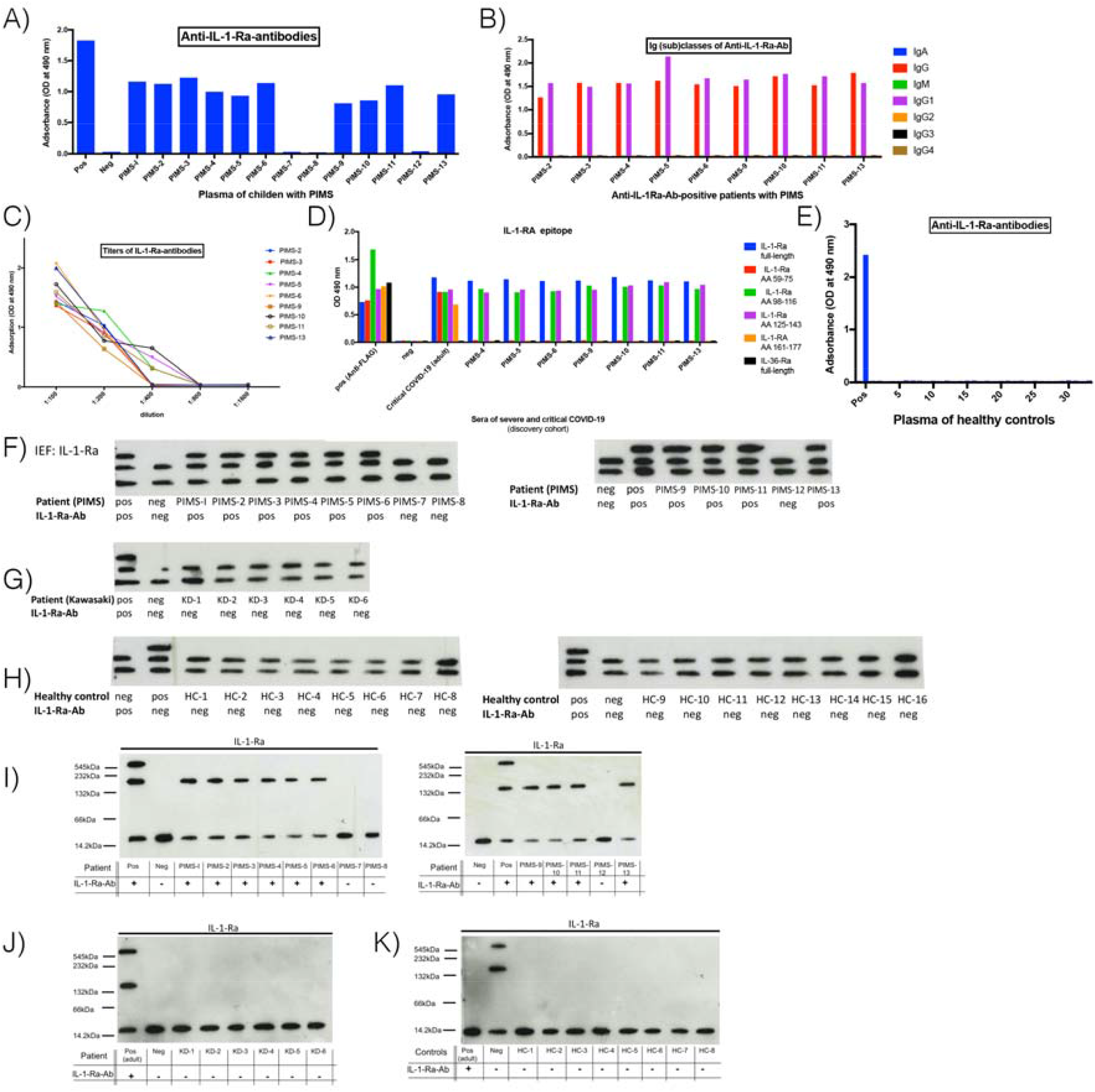
A) Occurrence of anti-IL-1-Ra-Abs in plasma from patients with PIMS/MIS-C. Anti-IL-1-Ra-Abs were found in 10 of 13 patients with PIMS/MIS-C. B) Ig classes and IgG subclasses of IL-1-Ra-Abs of the case-series. C) Titers of IL-1-Ra-Abs in patients of the case-series with PIMS/MIS-C. D) Epitope Mapping of IL-1-Ra-Abs in PIMS/MIS-C. E) Occurrence of anti-IL-1-Ra-Abs in plasma of healthy children F) IEF of IL-1-Ra in the patients with PIMS/MIS-C. An additional and differentially charged IL-1-Ra isoform appeared in IL-1-Ra-Ab-positive patients with PIMS/MIS-C, which was not detected in patients with Kawasaki disease G), or healthy controls H). I) Native Western-blots on band strength of IL-1-Ra in IL-1-Ra-Ab-positive or -negative patients with PIMS/MIS-C. IL-1-Ra-Ab-positive patients had reduced levels of free IL-1-Ra, but additional bands of IgG-bound IL-1-Ra. The positive control was plasma from an adult patient with critical COVID-19 showing IgG- and IgM-bound IL-1-Ra. Native Western-blots of plasma from patients with Kawasaki disease J), or of healthy controls K) did neither show reduced bands of free IL-1-Ra nor immunocomplexes of IL-1-Ra.

### ELISA and Western blot for plasma levels of IL-1-Ra

Using standard ELISA, we found that the plasma levels of free IL-1-Ra were strongly reduced in the index patient (PIMS-I) at the time of acute inflammatory syndrome (t0 (Nov 2020): 171.95 pg/ml), compared to 3 months later (t1 (Feb 2021): 1297.1 pg/ml) or 7 months later (t2 (Jun 2021): 847.5 pg/ml).

Native Western blot of total plasma protein produced a significantly weakened Il-1-Ra signal. Notably, though, an additional signal representing IgG-bound IL-1-Ra was found during acute MIS-C that was absent 7 month later, when the patient was seronegative for IL-1-Ra-Abs (Fig. 1 C). This reduced band of free IL-1-Ra in combination with IgG-bound IL-1-Ra was found in the case-series in every patient with IL-1-Ra-Abs, but in patients with Kawasaki disease or healthy controls (Fig. 2 I-K)

Regarding all 13 patients with PIMS/MIS-C, minimal IL-1-Ra plasma levels at acute inflammatory state were significantly decreased (Mann-Whitney test, two tailed: p = 0.007) in IL-1-Ra-Ab-positive patients (median 306.7 pg/ml, SD: 48.1 pg/ml), when compared to IL-1-Ra-Ab-negative cases (median 1845 pg/ml, SD: 247.9pg/ml).

Remarkably, in a patient PIMS-6 of the case series of whom a follow-up sample was available, the initial plasma sample was positive for IL-1-Ra-Abs and the hyperphosphorylated isoform of IL-1-Ra and the initial IL-1-Ra plasma level detected by ELISA was 323.49 pg/ml, whereas in the follow-up sample 5 weeks later lacked both IL-1-Ra-Abs and the hyperphosphorylated IL-1-Ra isoform (Fig 2F) and the IL-1-Ra plasma level had increased to 1642 pg/ml.

**Table 3:**
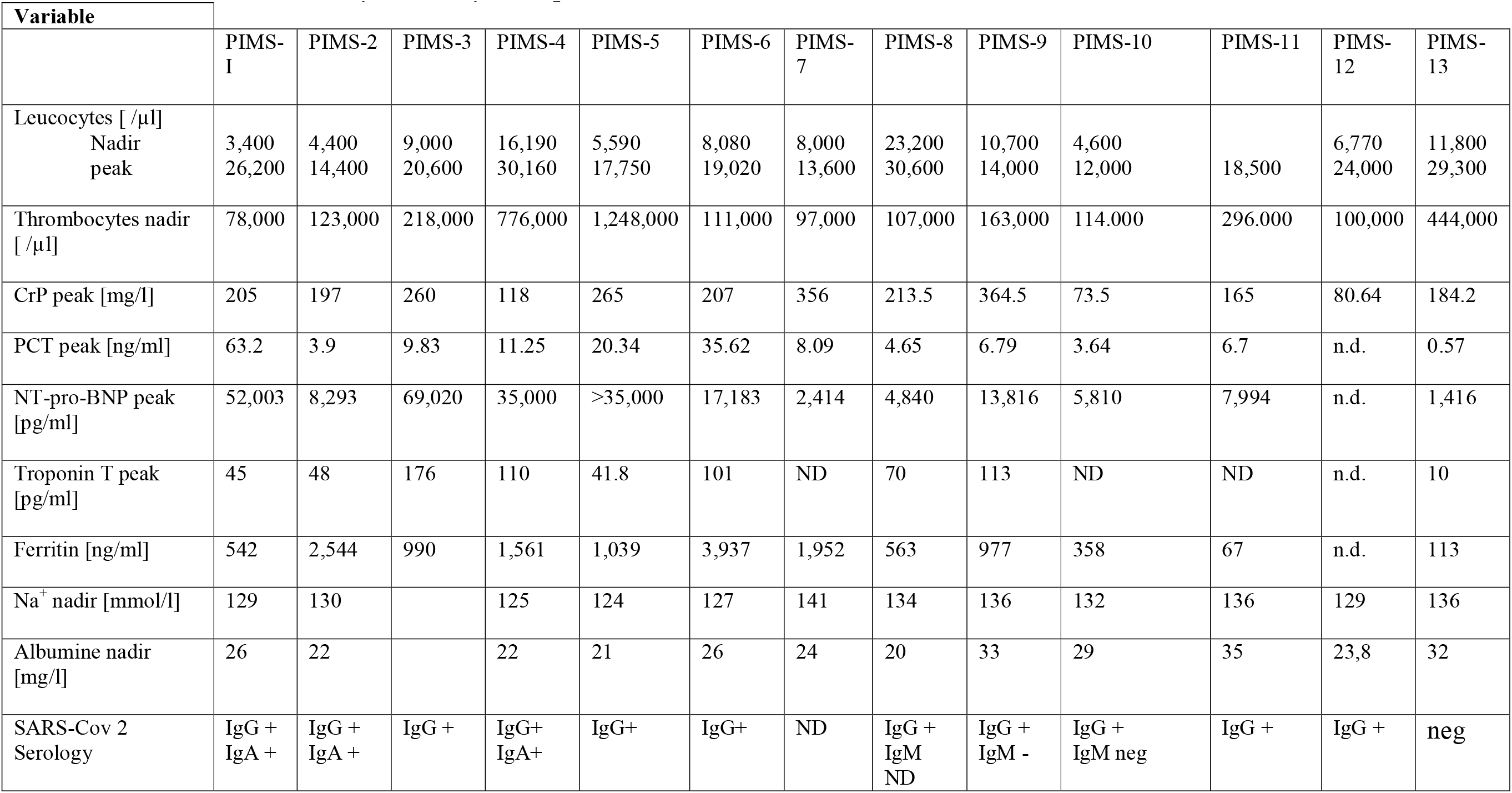
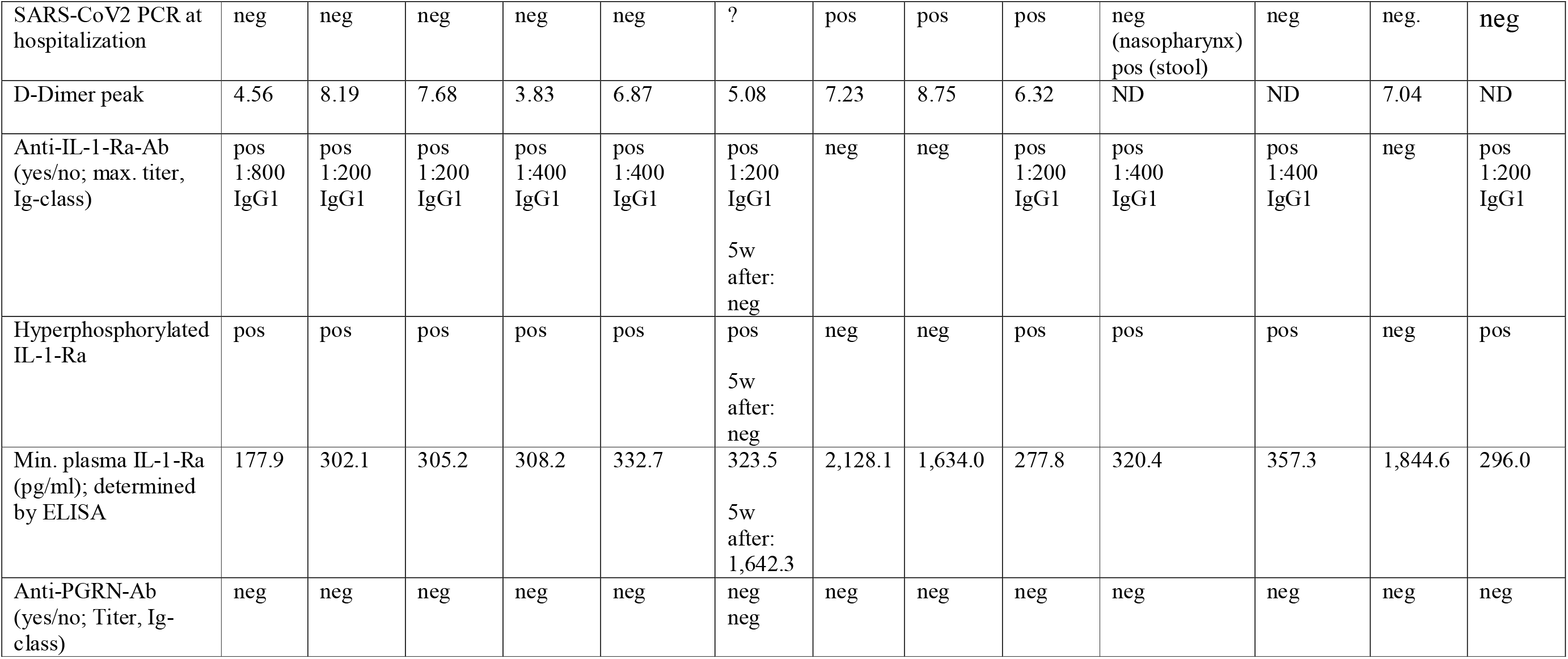
Clinical and laboratory summary of 13 patients with available clinical information.

### IL-1ß-Assay

Next, to investigate possible functional effects of the IL-1-Ra-Abs an in vitro IL-1ß assay was applied. Addition of plasma from patients with acute inflammatory syndrome and seropositive for IL-1-Ra-Abs significantly weakened the antagonism by recombinant IL-1-Ra, resulting in a stronger stimulatory effect of IL-1ß. The attenuation was comparable to the one seen upon the addition of a recombinant anti-human IL-1-Ra-antibody and to the addition of plasma from an adult patient with severe COVID-19 that had tested positive for IL-1-Ra-Abs. In contrast, though, addition of plasma from the same patient, at the same dilution, but 7 months after PIMS/MIS-C and lacking IL-1-Ra-Abs, did not weaken the effect of rec. IL-1-Ra (Fig. 3 B).

**Figure 3:**
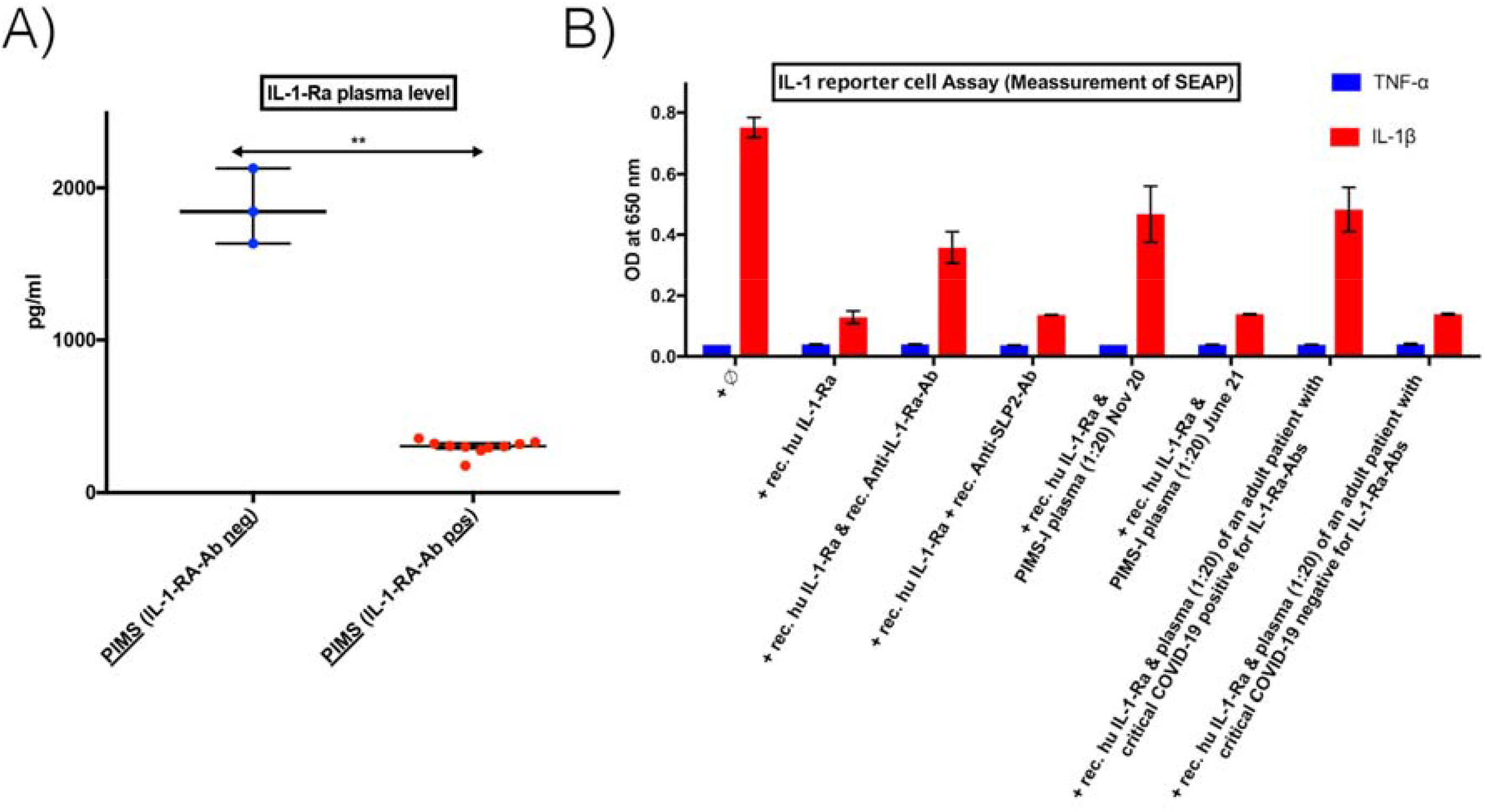
A) IL-1-Ra plasma levels determined by ELISA in patients with PIMS/MIS-C with IL-1-Ra-Abs (median 306.7 pg/ml, SD: 48.1 pg/ml), with PIMS/MIS-C without IL-1-Ra-Abs ((median 1845 pg/ml, SD: 247.9pg/ml), (Mann-Whitney test, two tailed: p = 0.007). Data are represented with median and interquartile range. \*\**P*≤0.01; \*\*\**P* ≤ 0.001; \*\*\*\**P*≤0.0001 B) Effect of IL-1-Ra-Ab-status on inhibition of IL-1ß. HEK IL-1 reporter cells were incubated with IL-1ß or TNF-α, rec. human IL-1-Ra, anti-IL-1-Ra-Ab, anti-SLP2-Ab as control, 1:20 diluted plasma of the index case (PIMS-I) during acute disease with IL-1-Ra-Abs, or 7 months after without IL-1-Ra-Abs, or 1:20 diluted plasma of IL-1-Ra-Ab-positive adult patient with acute critical COVID-19 or an adult IL-1-Ra-Ab-negative with critical COVID-19. The diluted plasma of the index case during acute inflammatory phase with IL-1-Ra-Abs resulted weakened the antagonistic effect of IL-1-Ra on IL-1ß. The adsorbance of SEAP, as a marker for IL-1 pathway activation in HEK IL-1 reporter cell was detected at 650 nm.

## DISCUSSION

Here we report on the high prevalence of neutralizing autoantibodies against the anti-inflammatory molecule IL-1-Ra in a case-series of patients with PIMS/MIS-C. Autoantibodies against IL-1-Ra were just recently described by our group in adults with critical COVID-19 (*14*); their occurrence was also reported in IgG4 associated diseases (*21*). In adults with critical COVID-19, IL-1-Ra-Abs were detected at a frequency of about 50%, along with autoantibodies against PGRN (in about 40% of patients). Moreover, in adults with critical COVID-19 we also found hyperphosphorylated isoforms of both IL-1-Ra and PGRN. All IL-1-Ra- and PGRN-Abs in adult COVID-19 patients belonged to IgM and different IgG subclasses.

In contrast to the findings in adult patients with critical COVID-19, no antibodies directed against PGRN were found in children with PIMS/MIS-C. IL-1-Ra-Abs were found at an even higher percentage in these children (10 of 13; 76,9%), compared to the adults with critical COVID-19. Notably, the plasma levels of free IL-1-Ra were strongly reduced in all patients with autoantibodies and simultaneously antibody-bound IL-1-Ra was detected. IL-1-Ra-Abs in PIMS/MIS-C belonged exclusively to IgG1 subclass, no antibodies of IgM class as in adults with critical COVID-19 were detected (*14*). The titers of IL-1-Ra-abs were lower compared to the adults with critical COVID-19 (1:200-1:800 in PIMS/MIS-C vs. 1:800-1:1600 in critical COVID-19 of adults).

A hyperphosphorylated isoform of the IL-1-Ra antigen was present in the plasma of all 10 patients with PIMS/MIS-C seropositive for IL-1-Ra-Abs. In the index patient, the disappearance of the hyperphosphorylated third isoform at follow-up preceded the disappearance of the IL-1-Ra-Abs. Longitudinal samples of adult patients with severe or critical COVID-19 had revealed that hyperphosphorylation of IL-1-Ra preceded the formation of autoantibodies. The data of COVID-19 of adults suggested that the immunogenic secondary modifications were induced by the SARS-CoV-2-infection itself or the inflammatory environment evoked by the infection (*14*), and that these modifications are immunogenic.

To further shed light on this issue, in-depth analysis of biobanked serum and plasma samples from children with complication-free courses after SARS-CoV-2 infection might be a next step. It should also prove useful to employ stimulation assays with various cytokines in order to trigger the hyperphosphorylated isoform of IL-1-Ra in established cell lines, primary cells of adults, and primary cells of children who i) had developed PIMS/MIS-C and ii) had a complication-free course after SARS-CoV-2 infection. Animal models have shown that low IL-1-Ra levels, either due to neutralization after immunization or due to deficiency, induce or exacerbate inflammatory syndromes such as colitis, arthropathy, vasculitis or psoriasis-like manifestations (*22*)(*23*)(*24*). In humans, inherited deficiency of IL-Ra (DIRA) results in a severe inflammatory syndrome (*25*)(*26*). In this context, autoantibodies against IL-1-Ra might, in part, explain the hyperinflammatory state in PIMS/MIS-C. The in-vitro results with plasma of patients with PIMS/MIS-C support this hypothesis.

IL-1-Ra-Abs did not occur in 6 cases of Kawasaki disease. Clearly though, further studies are needed to verify that antigen hyperphosphorylation and autoantibody presence are faithful discriminative markers. Moreover, the presented results point to strategies to strengthen current therapeutic approaches of IL-1-pathway inhibition and the standard therapy of IVIGs and glucocorticoids. Whether anakinra – as substitution for the decreased free IL-1-Ra levels but also as antigen in a preexisting autoimmune reaction - would make therapeutical sense or if a therapeutic IL-1ß antibody might be superior, needs also further exploration.

To summarize, autoantibodies against IL-1-Ra together with an immunogenic isoform of IL-1-Ra were observed in a PIMS/MIS-C case and subsequently in a high percentage of a case series. The relatively small number of cases included in the present study, due to the rarity of the disease, is a limitation. Nonetheless, these antibodies suggest to be pathogenetically relevant and should be further investigated in PIMS/MIS-C and other hyperinflammatory diseases.

## MATERIAL AND METHODS

This study was approved by the local Ethical Review Board (41/21) and conducted according to the Declaration of Helsinki. Blood Plasma samples of patients with PIMS/MIS-C according were taken after written informed consent in the department of Pediatric Cardiology of the Saarland University Hospital (Homburg/Saar, Germany), the Department of Pediatrics, Klinikum Saarbrücken (Germany), the Department of Pediatric Rheumatology and Immunology of the University Children’s Hospital Muenster (Germany), the Department of Pediatrics, Hospital Sant Joan de Déu, Universitat de Barcelona (Spain) and the Department of Pediatrics of Klinikum Kempten (Germany). All patients with PIMS/MIS-C fulfilled the WHO criteria, in addition all patients were seropositive for antibodies against SARS-CoV-2 or had positive PCR, with the exception of one patient who had only reported contact to SARS-CoV2.

In addition, 6 children with Kawasaki disease (age: min 1.4 years, max 7.5 years) and samples of 33 children with suspected growth retardation (age: min 3 years, max 15.3 years, median age 10.6 years) were included.

### ELISA for autoantibodies against PGRN, IL-1-Ra

The ELISA for autoantibodies was performed as previously described (*19*). In short, the antigens were obtained using the coding sequences of the *GRN* gene encoding PGRN, isoform 1 precursor of *IL1RN* were recombinantly expressed with a C-terminal FLAG-tag in HEK293 cells under the control of a cytomegalovirus promoter (pSFI). Total cell extracts were prepared and bound to Nunc MaxiSorp plates (eBioscience, Frankfurt, Germany) precoated with murine anti-FLAG mAb at a dilution of 1:2,500 (v/v; Sigma-Aldrich, Munich, Germany) at 4°C overnight. After blocking with 1.5% (w/v) gelatin in Tris-buffered saline (TBS) and washing steps with TBS with Triton X-100, the individual plasma samples were diluted 1:100. ELISA was performed according to standard protocols with the following Abs: biotinylated goat antihuman heavy and light chain immunoglobulin G (IgG) at a dilution of 1:2,500 (Dianova, Hamburg, Germany); subclass-specific sheep antihuman IgG1, IgG2, IgG3 and IgG4 (Binding Site Group, Birmingham, UK) at dilutions of 1:5,000; goat antihuman IgM (Dianova) at a dilution of 1:2,500; or goat antihuman IgA (Dianova) at a dilution of 1:2,500. Following this step, corresponding biotinylated secondary Abs were used for immunoassays carried out to detect IgG subclasses and IgM. Peroxidase-labelled streptavidin (Roche Applied Science, Indianapolis, IN, USA) was used at a dilution of 1:50,000. As a cut-off for positivity, the average of the optical density (OD) of the negative samples plus three standard deviations was applied. To narrow down the epitope region of the IL-1-Ra-Abs, full-length IL-1-Ra, following fragments thereof: amino acid 59-75, 98-116, 125-143 and 161-177, and as a control antigen full-length IL-36-Ra were recombinantly expressed with a C-terminal FLAG tag in HEK293 cells under the control of a cytomegalovirus promoter (pSFI).

### Western blot, isoelectric focusing of IL-1-Ra and PGRN

Isoelectric focusing (IEF) and Western blotting (including native Western blotting with non-reducing sample pretreatment and gradient gels without SDS) was performed. Plasma samples were analyzed for IL-1-Ra isoforms. Plasma from IL-1-Ra-Ab-positive patient was treated with alkaline phosphatase as previously described using FastAP thermo-sensitive alkaline phosphatase (Fermentas/VWR, Darmstadt, Germany) (*20*).

### ELISA for plasma level determination of IL-1-Ra

IL-1-Ra plasma levels were determined with a commercially available ELISA kit (Invitrogen/ThermoFisher #BMS2080) according to the manufacturer’s instructions.

### IL-1ß-assay

For IL-1ß assay HEK-Blue™ IL-1β reporter cells (Invivogen, #hkb-il1bv2) were used, which react specifically to IL-1ß and IL-1α by induction of NF-κB/AP-1, leading to expression of a secreted embryonic alkaline phosphatase (SEAP) reporter. Recombinant IL-1-Ra at 40ng/mL (Biozol, #PPT-AF-2000-01RA) alone or with either anti-IL-1-Ra antibody at 5 µg/mL (antibodies-online#ABIN2856394), recombinant SLP-antibody at 5µg/mL (abcam, #ab191883), plasma diluted 1:20 of a patient with acute PIMS/MIS-C and high-titered IL-1-Ra-antibodies (PIMS-I), or plasma diluted 1:20 of the same patient 7 months after PIMS/MIS-C without anymore detectable IL-1-Ra-Abs were preincubated for 2h at room temperature. Subsequently, these compounds were added with either IL-1ß (Biozol,# PPT-200-01B) or TNF-α (Biozol, #PPT-300-01A) both at 2ng/mL in 100µl DMEM to 2×10E4 HEK-Blue™ IL-1β reporter cells per well and incubated overnight at 37°C. Thereafter, 180 µL of each supernatant was transferred, 20µl QUANTI-BlueTM (Invivogen, #rep-qbs) was added and SEAP activity was measured at OD of 655nm. Experiments were performed in triplicate.

## Data Availability

Data can be provided upon request

## Author contribution

LTh, BTh, JP and CK planned the study, revised the data and wrote the manuscript. NF, ER, MCH and LTh performed experiments. KR, YF, KDP, SLB, MB, MB, CK, DF revised the data and manuscript. SM, SL, MCD, MF, HJ, HS, SM, TR, HAK, HW, KM, JA, RMP-R provided samples of patients and controls and clinical data.

## Acknowledgement

We thank the patients and their parents, who supported this research project. Further we want to thank all physicians, nurses and other staff not mentioned here, who cared for the patients. We thank Anja Geiger for proofreading of the manuscript. This work was supported by a young investigator NanoBioMed fund of the University of Saarland to LTh.

